# Performance evaluation of a non-invasive one-step multiplex RT-qPCR assay for detection of SARS-CoV-2 direct from human saliva

**DOI:** 10.1101/2022.03.09.22272125

**Authors:** Harry H. Jenkins, Ana A. Tellechea Lopez, Francesco Saverio Tarantini, Hannah Tomlin, Danielle Scales, I-Ning Lee, Siyu Wu, Ralph Hyde, Katarzyna Lis-Slimak, Timothy Byaruhanga, Jamie L. Thompson, Sara Pijuan-Galito, Lara Doolan, Kazuyo Kaneko, Penny Gwynne, Caroline Reffin, Emily Park, Jayasree Dey, Jack Hill, Asta Arendt-Tranholm, Amy Stroud, Moira Petrie, Chris Denning, Andrew V. Benest, Claire Seedhouse

**Affiliations:** University of Nottingham Asymptomatic Testing Service (UoNATS); Biodiscovery Institute, School of Medicine, University of Nottingham, University Park, Nottingham, England, NG7 2RD

## Abstract

Polymerase chain reaction (PCR) has proven to be the gold-standard for SARS-CoV-2 detection in clinical settings. The most common approaches rely on nasopharyngeal specimens obtained from swabs, followed by RNA extraction, reverse transcription and quantitative PCR. Although swab-based PCR is sensitive, swabbing is invasive and unpleasant to administer, reducing patient compliance for regular testing and resulting in an increased risk of improper sample collection. To overcome these obstacles, we developed a non-invasive one-step RT-qPCR assay performed directly on saliva specimens. The University of Nottingham Asymptomatic Testing Service (UoNATS) protocol simplifies sample collection and bypasses the need for RNA extraction, additives, or extraneous processing steps, thus helping to encourage more regular testing and reducing processing time and costs. We have evaluated the assay against the performance criteria specified by the UK regulatory bodies and attained accreditation (BS EN ISO/IEC 17025:2017) for SARS-CoV-2 diagnostic testing by the United Kingdom Accreditation Service (UKAS). We observed a sensitivity of 1 viral copy per microlitre of saliva and demonstrated a concordance of >99.4% between our results and those of other accredited testing facilities. We concluded that saliva is a stable medium with surprising longevity, and allows for a highly precise, repeatable, and robust testing method.

## Introduction

Since the emergence of Severe Acute Respiratory Syndrome Coronavirus 2 (SARS-CoV-2) in 2019 in the Wuhan province China ^[1,2]^, much attention has been given to the necessity of surveillance and molecular testing amongst populations worldwide to curb viral propagation. SARS-CoV-2 is the third known coronavirus capable of animal-to-human transmission, after SARS-CoV and MERS-CoV ^[2]^. However, SARS-CoV-2 is a greater issue than its predecessors in terms of the number of global infections and infection-related mortality ^[2]^ and has fast become a global pandemic.

As of the 1st of February 2022, UK government statistics reported over 17 million confirmed COVID-19 cases since the emergence of the pandemic, resulting in over 176,000 deaths. Globally, the World Health Organisation (WHO) statistics estimate over 373 million confirmed cases with over 5.6 million deaths. Indeed, public health services have endured immense pressure amid rising hospital admissions, imparting considerable strain on an already overstretched medical staff. The majority of SARS-CoV-2 diagnostic testing is performed only when an infection is suspected, likely following the onset of characteristic symptoms e.g., fever, persistent cough, or the loss of taste or smell. However, there is now a greater appreciation for the impact of infections that are asymptomatic or have non-standard characteristics on subsequent transmission through communities. Indeed, meta-analysis shows that approximately 30% of infections remain asymptomatic ^[3,4]^. Evidence suggests that general population testing can reduce transmission, and is correlated to a decline in SARS-CoV-2-related mortality ^[5]^.

The frontline and current gold-standard diagnostic tests for the detection of SARS-CoV-2 infection utilise nucleic acid amplification tests usually via RT-qPCR, most commonly from specimens obtained through nasopharyngeal swab (NPS) or oropharyngeal swab (OPS). Swab testing, although commonplace, is highly dependent on correct administration to yield accurate results; however, it is considered invasive and unpleasant by many, unsuitable for the very young or some individuals with learning disabilities, and the discomfort induced during an accurate swabbing procedure may potentially result in an unwillingness to test regularly ^[6]^. Importantly, inappropriate nasopharyngeal swab sampling may associate with false-negative results, thus contributing to the spread of infection to the general population ^[7]^, and reducing confidence in the accuracy of the tests. SARS-CoV-2 diagnostic tests utilising saliva therefore present an attractive alternative whilst being cheaper to process ^[8,9]^.

Saliva is undoubtably a very heterogenous substance comprising electrolytes, proteins, lipids, amino acids, gingival crevicular fluid, antibodies, immune and epithelial cells, food matter, bacteria, and viruses ^[10,11]^. Nonetheless, the current literature contains a wealth of evidence in support of the use of saliva as a suitable medium for some clinical diagnostic applications. To *et al*. report that salivary viral load is highest in the first week of infection, and the virus detectable in saliva up to 25 days after the onset of symptoms ^[12]^. Furthermore, as angiotensin converting enzyme 2 (ACE-2), the entry receptor for SARS-CoV-2, is expressed within the oral mucosal epithelial cells ^[13,14]^, and oral fluids have the potential to transmit SARS-CoV-2 infection ^[9]^, the concept that saliva is a key site for viral detection is supported. A study by Silva *et al*. has shown that the salivary viral load correlates to COVID-19 disease severity and to poor prognosis ^[9,15]^. Willie *et al*. report that saliva specimens provided a greater sensitivity for viral detection than swabs from hospital in-patients ^[16]^. A meta-analysis by Czumble *et al*. has found the sensitivity of saliva tests to be slightly lower yet comparable to those based on NPS (91% c.f. 98% respectively) ^[10]^. However, the inconsistencies in the administration of the swabs and the specific PCR protocols used to diagnose the infection have to be considered for this comparison.

The impact that the SARS-CoV-2 pandemic has had on the provision of teaching and the continuation of research in higher education institutions was devastating. At the University of Nottingham, we identified the need to establish a SARS-CoV-2 testing service to be offered to all university students and staff in order to monitor transmission within our community, hence allowing mitigating actions to reduce further transmission chains. To this end, we developed a simple, yet sensitive multiplexed RT-qPCR assay intended for the diagnostic detection of SARS-CoV-2 RNA in heat-inactivated saliva samples, specifically in asymptomatic individuals. The use of saliva bypasses the need for swab-based specimen provision and allows for effortless self-sampling, thus minimising the risk of improper administration and reducing the requirement for consumables in national shortage (such as swabs). Furthermore, our one-step assay eliminates the time-consuming and costly RNA purification steps and is based on the Centre for Disease Control and Prevention (CDC) approved Quantabio^®^ UltraPlex 1-Step ToughMix™, an RT-qPCR reaction mix developed to be resistant to PCR inhibitors that can be present in crude biological samples such as saliva ^[17]^.

The detection of viral genome in the sample relies on three sets of primers and probes specific for as many gene targets (*Table 1*): The envelope (E) gene (from the Charité/Berlin method, as listed in the WHO-approved SARS-CoV-2 detection protocols), the nucleocapsid (N) gene (from the US-CDC’s nCOV_N2 assay), and the human RNase P gene (used as a control for saliva-sample integrity in the US-CDC_RNase P assay) ^[17-19]^.

**Table 1:**
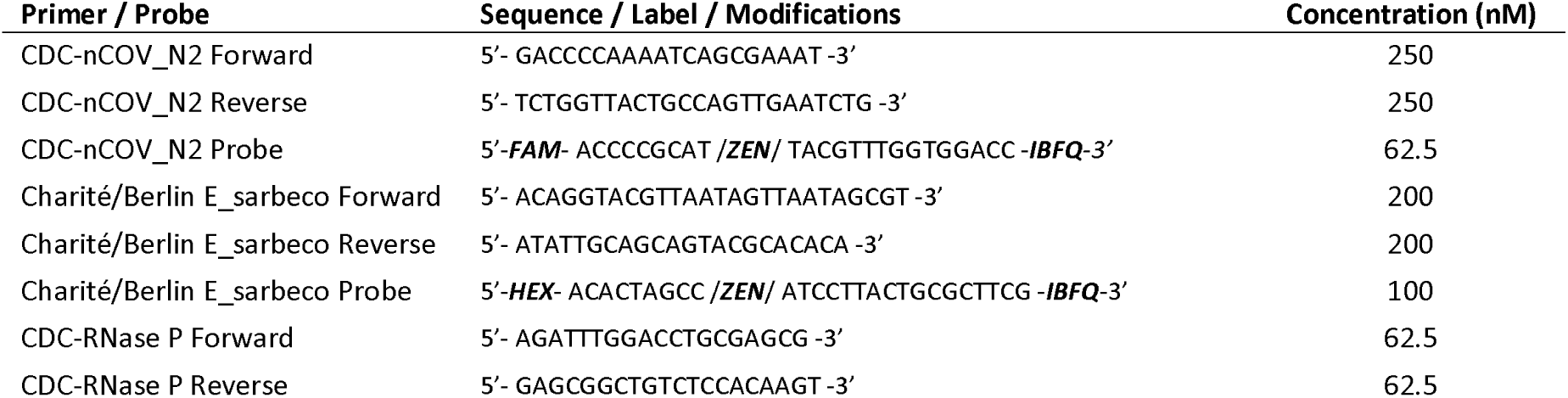
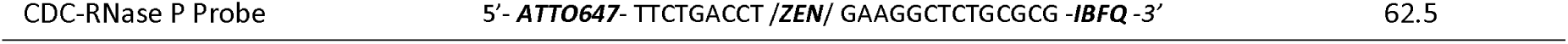
Primers and probes used in the assay, with respective sequences, modifications, and final qPCR concentrations.

In 2021, the UoNATS was assessed by the United Kingdom Accreditation Service (UKAS), and subsequently awarded accreditation as a SARS-CoV-2 testing provider (Testing Laboratory No. 22743). The following experimental data describe the methods used for the in-house validation of the UoNATS direct RT-qPCR diagnostic test, as well as a comparison with results generated from independent accredited laboratories. The validation work was performed with consideration to the Public Health England’s (PHE) published quality guidance on the UK Standards for Microbiology Investigations (UK SMI) ^[20]^, the Medicines and Healthcare Regulatory Agency (MHRA) target product profile (TPP) specifications for SARS-CoV-2 viral detection testing, and adhering to the applicable criteria defined within the European Standard for the competence of testing and calibration laboratories (BS EN ISO/IEC 17025:2017). Whilst a parallel submitted manuscript (Tarantini *et al*., 2022 ^[21]^) provides stepwise protocols for the UoNATS assay, the article presented below defines the quality metrics used to validate and accredit the UoNATS SARS-CoV-2 detection method, as assessed by the following parameters: Linearity and range, analytical sensitivity (limit of detection, LOD), analytical specificity, diagnostic (clinical) sensitivity and specificity, positive and negative predictive value, repeatability and sample stability, accuracy and precision, multiplex validation, robustness and reproducibility.

## Materials & Methods

### Ethical Approval

The studies involving human participants were reviewed and approved by University of Nottingham Ethics Committee, reference number FMHS 96-0920. The patients/participants provided informed consent to participate in this study.

### Specimen collection and processing

Detailed methods have been provided in a separate publication ^[21]^. Clinical samples are inactivated prior to downstream processing; vials are heat-inactivated in a laboratory oven with the use of a temperature probe to ensure samples are held at 95 °C for 5 minutes, shown to inactivate SARS-CoV-2 ^[22]^.

### RT-qPCR

RT-qPCR is performed in a one-step protocol using primer and probe sequences as recommended by the US-CDC and the Charité/Berlin panel ^[18]^; US-CDC-nCOV_N2:FAM, US-CDC_RNaseP:ATTO647, Charité-E:HEX (Integrated DNA Technologies). Quantabio UltraPlex™ 4x Toughmix (VWR; Cat. 95166) mastermix was used. Oligo sequences and final concentrations are given in (*Table 1*); reagents used in each 20 µL qPCR reaction were 5 µL of Ultraplex™ ToughMix (4x), 0.25 µL of primer and probe mix, 6.75 µL of nuclease-free water and 8 µL of heat-inactivated saliva. Nuclease-free water, and commercial SARS-CoV-2 standard at a known concentration of 2 viral copies per microlitre (vc µL^-1^) are included in triplicate in every run as non-template and positive controls, respectively. All qPCR reactions were performed using 96-well low-profile PCR plates (Eppendorf TwinTec; VWR Cat. No. 732-0107) and optical plate sealers (Thermo-Fisher; Cat. No. AB-0558), on a Bio-Rad C1000 thermal cycler with CFX96 Touch™ optical unit, cycling conditions are described in (*Table 2*). Data analysis is performed using the Bio-Rad CFX Maestro software package. Positivity is determined where the CT for 1 or both SARS-CoV-2 targets is <35; an inconclusive result is attributed to a sample with CT of >35 but <37; results of CT >37 are deemed negative.

**Table 2:** RT-qPCR Cycling conditions. Acquisition of FAM: 510-530nm; HEX: 560-580nm; ATTO647 (Cy5 channel): 675-690nm.

| Step | Temperature (°C) | Duration (min:sec) | Cycles |
| --- | --- | --- | --- |
| Reverse Transcription | 50 | 10:00 | x1 |
| RT Inactivation/Initial Denaturation | 95 | 03:00 | x1 |
| Denaturation | 95 | 00:03 | x45 |
| Annealing | 55 | 00:30 |  |
| Extension (Acquisition) | 72 | 00:15 |  |

### Linearity and Range

To assess the linearity and dynamic range of the assay, a previously tested clinical SARS-CoV-2-positive sample was serially diluted in a pool of SARS-CoV-2-negative saliva derived from 30 individual negative samples and tested in triplicate. R^2^ values of >0.95 are considered acceptable. The CT range generated spans 15-40 which is considered representative of high-to-low viral loads and comprises the vast majority of samples received at our facility.

### Analytical Sensitivity (Limit of detection, LOD)

A commercial external run control (NATrol™ NATSARS(COV2)-ERC, ZeptoMetrix Corp.) consisting of whole chemically-inactivated virions at 50 vc µL^-1^ was diluted from 1:3.125 (16 vc µL^-1^) to 1:400 (0.125 vc µL^-1^) in serial two-fold dilutions using a pool of combined negative saliva from 30 previously tested SARS-CoV-2-negative samples as the diluent. Replicate samples (n=6, or n=12 for lower concentrations) were tested. The experimental setup was performed in duplicate by separate operators using different pools of negative saliva diluent. The limit of detection is defined as the lowest concentration tested whereby 100% of replicates are detected by the assay.

### Analytical Specificity: Cross reactivity of the UoNATS reaction mix

To assess potential cross-reactivity of the UoNATS assay with other commonly infecting pathogens, a commercial respiratory verification panel (NATRVP2.1-BIO; ZeptoMetrix, USA) was used as a control. The panel contains 21 inactivated microorganisms known to infect the respiratory system of humans, thus with the potential to be present in the specimens collected in the UoNATS workflow.

To monitor the on-going ability of the assay to detect prevalent variant strains of SARS-CoV-2, the primer probe sequences are periodically BLAST-aligned against sequences submitted to the EpiCov database, hosted by the Global Initiative for Sharing All Influenza Data (GISAID). Strains with 2 or more mutations in each of the assay target regions, or 1 or more mutation within the 3’ end of each target sequence are reported.

### Diagnostic Sensitivity and Diagnostic Specificity

The diagnostic (clinical) sensitivity and specificity was assessed by testing both positive (n=163) and negative (n=250) saliva specimens and nasopharyngeal swab specimens submitted by the same donor preferably on the same day. Exclusion criteria were if swab and saliva had been provided >5 days apart. The swab samples were tested by independent accredited testing facilities: Source Bioscience; Nottingham, UK (Laboratory No. 9571) & Nottingham University Hospitals NHS Trust (Laboratory No. 8848), saliva samples were tested in-house. Clinical concordance of the UoNATS results with those of external testing facilities were calculated as the percent agreement of the results. Concordance of 95% to 100% is considered acceptable.

### Positive and Negative Predictive Values (PPV and NPV)

To estimate the risk of false-positive or false-negative results, the diagnostic sensitivity and specificity were each used to calculate the PPV and NPV, respectively for the UoNATS test. PPV is calculated using the formula: PPV = true positives/(true-positives + true negatives) and NPV is calculated using the formula: NPV = true negatives/(false negatives + true negatives), as specified by Public Health England (PHE) ^[20]^. The term ‘true’ herein shall refer to the results derived from independent testing facilities (Source Bioscience; Nottingham, UK & Nottingham University Hospitals NHS Trust) using a gold-standard (i.e., RT-qPCR) testing methodology for the reference method. The positive and negative predictive values were then estimated for several scenarios with low to high prevalence in the population (1, 5, 10, 25 and 50%) as specified by the MHRA TPP for laboratory-based SARS-CoV-2 viral detection tests (version 2.0), and for the estimated prevalence for November 2021 for the University of Nottingham students and staff population (0.61%) as determined by our own workflow. NPV and PPV were determined for specimens tested a single time as first-line screening, and after a subsequent second confirmatory test.

### Sample Stability, Repeatability, Reproducibility

To ascertain the stability of viral nucleic acid in saliva, three SARS-CoV-2-positive samples were aliquoted on the day of detection and aliquots (n=10) stored at room temperature (18-22 °C), 4 °C, or -80 °C. Aliquots kept at -80 °C were re-usable and so represent a sample with multiple freeze/thaw cycles. Each set of aliquots was tested in triplicate every 2-3 days for a period of 20 days. Stability of viral RNA in the medium was inferred by the consistency of CT values throughout the time course.

The repeatability of the assay was determined by analysing the mean CT, standard deviation, and co-efficient of variation derived from the daily use of a standard of known concentration (2x LOD) over 100 days; the co-efficient of variation should not exceed 5%.

To establish the reproducibility of results, 7 SARS-CoV-2-positive samples (including 1 known problematic specimen; sample 7) were tested in triplicate by each one of four laboratory operators using independently prepared reaction mixes and equipment. Tests were performed on the same day to exclude CT variations linked to storage conditions.

### Accuracy and Precision

An independently validated negative sample was spiked with an external positive control (NATrol™ NATSARS(COV2)-ERC, ZeptoMetrix Corp.) consisting of whole chemically inactivated virions at the limit of detection for the assay (determined as 1 vc µL^-1^). The sample was tested in 24 technical replicates by the same operator, standard deviations and co-efficient of variation was calculated; %CV should not exceed 10% for confidence in the accuracy and precision of the method.

### Multiplexing Validation

To assess if multiplexing the three primer probe sets resulted in any inhibition to the reactions which may affect the quantification and subsequent test result, dilutions of a previously tested SARS-CoV-2 positive clinical sample were produced and tested using; 1) CDC-N2 only; 2) Charité-E only; 3) UoNATS triplex N2/E/RP assay conditions. CT changes between mono/triplex of ≤ 1 were considered acceptable.

### Robustness

The robustness evaluation of the assay was determined by quantifying the variation in results derived from the assay when under pressure from confounding variables; including, inaccurate pipetting volumes, and the uncontrollable presence of PCR inhibitory factors which may be present in the heterogenous saliva matrices. Co-efficient of variations of 0-5% were considered acceptable.

## Results

### Linearity and Range

To establish the accuracy of the assay over a range of viral concentrations, a strong positive clinical sample was serially diluted to yield a CT range of 15 – 40, encompassing the expected range for the vast majority of the samples tested in our workflow and intended to represent high-to-low viral load. PCR efficiencies for both N2 and E primer/probe assays were calculated at 107% when combined in the multiplexed assay, with R^2^ values all >0.98 demonstrating accuracy and precision through this range (*Fig. 1a*).

**Figure 1:**
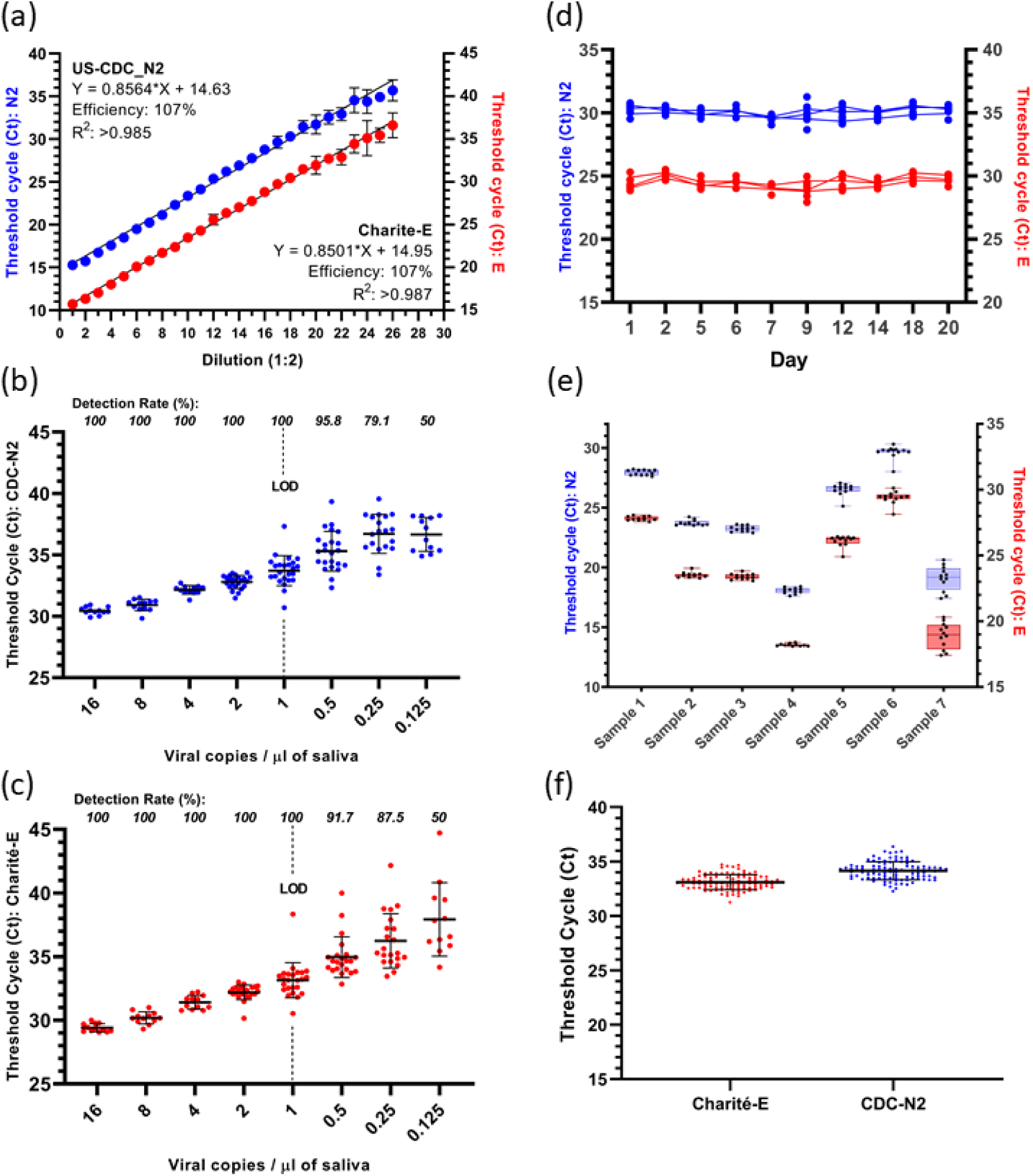
Data in blue represents the CDC-N2 assay, and red data for Charité-E. Mean CT is plotted, error bars denote the standard deviation. **(a)** PCR efficiency and linear range derived from a serially diluted known positive clinical sample, spanning a range of high-to-low viral loads. The detection rates and LOD for the N2 and E assays are shown in panels **(b)** and **(c)** respectively. **(d)** The CT values of a positive sample tested over 20 days under various storage conditions. **(e)** Robustness results between operators and reagents. **(f)** Repeatability and reproducibility when sequentially testing a commercial standard at near-LOD daily for 100 days.

### Analytical Sensitivity (Limit of detection, LOD)

The limit of detection (LOD) of the assay was determined as the lowest viral concentration at which 100% detection is observed. A requirement for tests to detect viral loads of ≤1 vc µL^-1^ are specified by the relevant UK governing bodies (PHE & MHRA). The UoNATS assay can reliably detect 100% of replicates with a viral load equivalent to 1 viral copy per microlitre (vc µL^-1^) of saliva or above, for both gene targets. *Figure 1b & 1c* present the results from viral concentrations between 16 vc µL^-1^ to 0.25 vc µL^-1^. Each plot shows the combined results from 2 separate experiments.

### Analytical Specificity

To ensure the UoNATS assay does not detect other respiratory-infecting microorganisms which may be present in deposited samples, we tested the assay using a commercial respiratory verification panel consisting of 21 respiratory pathogens. There was no reactivity with either of the SARS-CoV-2 targets, N2 or E. A positive result was determined for both targets for SARS-CoV-2 (*Supplementary Table S2*).

Primer and probe sequences used in the UoNATS assay were BLAST-aligned against submitted SARS-CoV-2 sequences in the EpiCov (GISAID) database to ensure the assay will confidently detect variants of concern. Those strains with 2 or more mutations in the assay regions, or 1 mutation if in the 3’ end, were quantified (*Supplementary Table S3*). Omicron (B.1.1.529) and sub lineages BA.1, BA.1.1, BA.2, and BA.3, now >95% of sequences in Europe (EpiCov; GISAID) have a single C>T mismatch in the 5’ end of the Charité-E forward primer binding region, this has not been observed to affect assay performance. Only 1 of 495 strains analysed (0.2%) had a single mismatch in a 3’ assay region, from an isolate of the BA.2 sub lineage. No prevalent variants were identified with mutations in both N2, and E target regions simultaneously thus likely to affect viral detection in our assay.

### Diagnostic Sensitivity & Specificity

The clinical sensitivity and specificity of the method was assessed by comparing the results from SARS-CoV-2 nasopharyngeal swabs tested by independent accredited testing facilities (Source Bioscience, Nottingham, UK; and Nottingham University Hospitals NHS Trust) with UoNATS saliva results. The concordance was calculated by matching the results of 250 samples deemed negative by external testing facilities using our assay, all confirmed with exception of one false positive; and 163 samples tested positive by external testing facilities, also confirmed using our assay with one exception, being detected but falling beyond our internal CT cut-off of 37. We observed a 99.4% and 99.6% agreement with external positive and negative results, respectively, satisfying the acceptance criteria (>95%).

### Positive (PPV) and Negative (NPV) Predictive Values for various SARS-CoV-2 prevalence

To determine the number of false positives and false negatives which this assay could generate at fluctuating SARS-CoV-2 prevalence, a population of 10,000 was considered and the clinical sensitivity and specificity used. For a first round of tests, initial screening, the PPV was between 60-100% and the NPV was between 99-100%. When the number of positive samples detected are subjected to a second test, confirmatory test, the overall PPV is 99-100% and the NPV, 100%. Details of estimated predictive values based on populations with varying SARS-CoV-2 incidence are given in *Table 3* (and described in *Supplementary Table S7*). In each case, the positive predictive value (PPV) was obtained by dividing the number of true positive cases into the total positive results. The negative predictive values (NPV) by dividing the number of true negative cases into the total negative results (see detailed methods).

**Table 3:**
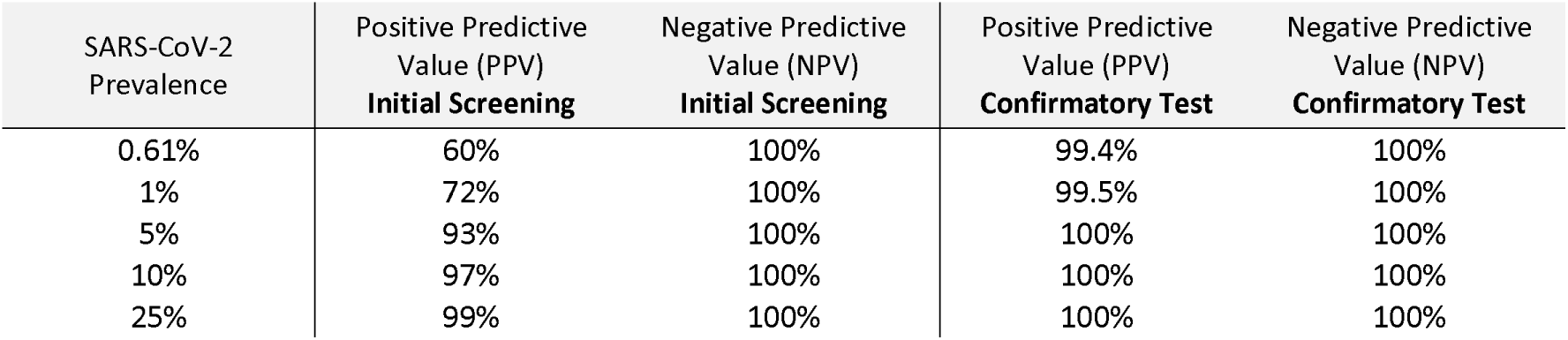
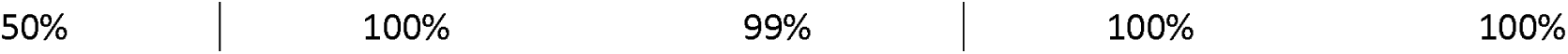
Positive and Negative predictive value calculations after initial screening and confirmatory test of positive samples.

### Sample Stability, Repeatability, and Reproducibility

To further investigate the performance of the assay, we questioned the stability of viral nucleic acid in saliva over time in different storage conditions. SARS-CoV-2 RNA in clinical samples was stable within saliva for up to 20 days irrespective of storage temperature (RT: 18-22 °C, 4 °C, or -80 °C), and was robust through at least 8 freeze/thaw cycles without detriment to the CT values obtained (*Fig. 1d*). Detailed statistics are given in *Supplementary Table S4*.

The mean CT values from daily testing of a commercial standard over a period of 100 days were collated and used to quantify the repeatability and reproducibility of the assay. This test reflects multiple batches of reagents, alternative users, PCR machines, and pipettes. Our assay demonstrated highly repeatable results, with co-efficient of variations of <2.4% and <2.1% for N2 and E, respectively (*Fig. 1f*).

We next tested 7 clinical positive samples using different PCR machines, reagents, and laboratory members to estimate the reproducibility of the method. We observed a maximum co-efficient of variation of <2.4%, with the exception of 1 sample (sample 7, 5.4% CV) which was highly viscous and difficult to process (*Fig. 1e*).

### Accuracy and Precision

The descriptive statistics determined from multiple replicate tests of the same sample are presented in *Supplementary Table S5*. Commercially sourced SARS-CoV-2 positive run control was added into SARS-CoV-2 negative saliva to a concentration of 1 vc µL^-1^ corresponding to the limit of detection of the assay where variability would be most pronounced. A standard deviation of 0.9 and 1.0 and co-efficient of variation of 2.6% and 2.9% were observed across individual replicates (n=24) for the N2 and E targets, respectively, demonstrating good precision of measurement.

### Multiplex Validation

To ascertain if multiplexing the primer/probe assays is detrimental to the PCR performance, serially diluted SARS-CoV-2 positive saliva was tested; the CT values for the individual N2, or E reactions were compared to those derived from the same samples using the multiplexed reaction mixture. The results showed a high concordance (see *Supplementary Fig. S1*) with mean CT differences between mono-and triplex of 0.25 for N2, and 0.17 for E.

### Robustness

#### Influence of matrix in the assay

To address the question of the heterogeneity of saliva on the accuracy of the assay; 28 samples were randomly selected from a pool of clinical samples previously confirmed as negative. Viral particles from commercial standards were added to each sample to a concentration of 1 vc µL^-1^ (LOD). Each sample was tested and the mean CT ± S.D., and coefficient of variation (%CV) were calculated. The CT values did not deviate markedly and %CV was <3.5% suggesting that varying compositions of saliva do not inhibit the ability to detect SARS-CoV-2 precisely (*Table 4*).

**Table 4.** Quantifying variability from heterogeneous saliva matrices on the performance of the UoNATS assay; 28 different saliva samples were spiked with reference standard at the LOD of 1 vc µL^-1^ to measure the effect of uncontrollable inhibitory factors on the accuracy of results.

|  | 1x LOD (CDC-N2) | 1x LOD (Charité-E) |
| --- | --- | --- |
| Number of samples | 28 | 28 |
| Mean CT | 34.54 | 34.89 |
| Std. Deviation | 1.189 | 0.920 |
| Coefficient of variation | 3.442% | 2.637% |

##### Influence of pipetting volumes in the assay

To assess the effect of potential pipetting inaccuracies (due to sample viscosity) an external positive control was added to negative saliva to a concentration of 2 vc µL^-1^ (2x LOD) and added in triplicate as 6, 7, 8, 9 and 10 µL sample volumes to 12 µL of master mix. The effects are summarised in *Table 5* and demonstrates a low variability in CT values as a result of pipetting deviations of ± 2 µL from an 8 µL sample volume.

**Table 5:** Descriptive statistics showing the effect of controlled variation of volumes of saliva in a 20 µl reaction.

| CDC-N2 (FAM) |  |  |  |  |  |
| --- | --- | --- | --- | --- | --- |
| | 6 $\mu\text{L}$ | 7 $\mu\text{L}$ | 8 $\mu\text{L}$ | 9 $\mu\text{L}$ | 10 $\mu\text{L}$ |
| Number of replicates | 3 | 3 | 3 | 3 | 3 |
| Mean CT | 33.38 | 33.61 | 32.92 | 33.72 | 34.25 |
| Std. Deviation | 0.227 | 0.612 | 0.921 | 1.505 | 0.302 |
| Coefficient of variation | 0.680% | 1.821% | 2.796% | 4.463% | 0.882% |

| Charité-E (HEX) |  |  |  |  |  |
| --- | --- | --- | --- | --- | --- |
| | 6 $\mu\text{L}$ | 7 $\mu\text{L}$ | 8 $\mu\text{L}$ | 9 $\mu\text{L}$ | 10 $\mu\text{L}$ |
| Number of replicates | 3 | 3 | 3 | 3 | 3 |
| Mean CT | 32.25 | 32.77 | 32.00 | 32.40 | 33.70 |
| Std. Deviation | 0.106 | 0.659 | 0.609 | 1.228 | 0.475 |
| Coefficient of variation | 0.329% | 2.011% | 1.902% | 3.791% | 1.408% |

##### Problematic samples

Saliva specimens with a highly viscous and gel-like consistency are difficult to pipette accurately. A clinical specimen of this type was analysed to ascertain potential inaccuracies in the results which may be expected with such samples. Four operators tested the same sample in triplicate, *Supplementary Table S6 (Sample 7)* summarises the results for each gene. Although this sample represents one of the worst examples of saliva to pipette, the coefficient of variation for N2 and E were lower than 5.4% when tested with the UoNATS assay.

## Discussion

The SARS-CoV-2 pandemic created an unprecedented need for regular viral population-based testing. Of the solutions that have been utilised for this purpose, PCR-based diagnostic tests have constituted the frontline method of choice. Rapid tests have been developed recently to attempt to combat bottlenecks in the national and international testing strategies; lateral flow devices (LFD) to detect SARS-CoV-2 antigens have become widely used despite concerns around their accuracy; a Cochrane systematic review determined that the sensitivity of antigen tests range from as low as 34.1% to 88.1% ^[23]^. The variable sensitivity of LFD devices will tend to result in a proportion of asymptomatic infections going undetected. This is especially true for those people with lower viral loads including vaccinated individuals ^[24]^ and hence will likely increase in line with uptake of vaccination. Furthermore, as SARS-CoV-2 variants emerge, there may be a selective pressure favouring the propagation of less virulent strains.

Most of the currently available techniques, both antigen-based and nucleic acid-based, rely on nasopharyngeal swabs for specimen collection. Accurate diagnostic testing ultimately relies on accurate self-administration of swabs; many people find this unpleasant, and this increases the likelihood of ineffective sampling and potentially false-negative results ^[6,7]^. Alternative non-invasive methods of sampling will prove essential if frequent mass testing of individuals can be maintained both logistically and financially long-term throughout this pandemic. Using saliva overcomes the unwillingness of donors to administer unpleasant swabs, which may encourage more regular testing, and therefore detect cases which would otherwise be undiagnosed.

The University of Nottingham’s SARS-CoV-2 testing service was established in August 2020 to offer regular asymptomatic PCR testing to all students and staff, free-of-charge, in a bid to curtail transmission chains. We have since extended this service to the young children of university affiliates. The service enabled the university to maintain a safe environment allowing a return to campus, and the continuation of uninterrupted research and face-to-face teaching. As of February 2022, we have processed in excess of 130,000 samples. Of these, there were >1,300 SARS-CoV-2 positive cases in individuals who were classified as asymptomatic by clinical guidance at the time. Since these were not eligible for PCR testing through government and national health channels, this shows direct evidence for decreasing the likelihood of transmission through the community. Of course, such a testing regime incurs a substantial financial burden; however streamlined and cost-effective testing methodology has allowed long-term preservation of the service whilst maintaining highly sensitive and reproducible results.

Some debate in the literature has arisen as to the sensitivity and accuracy of alternative (non-swab based) sample media. Saliva specimens are considered variable by some; however, others show that both viral stability and assay sensitivity using saliva can meet comparable specifications to that of correctly administered NPS ^[10]^. We sought here to reinforce the idea that saliva can provide a reliable and highly sensitive medium for the detection of SARS-CoV-2 nucleic acid and should be utilised to a greater extent. Furthermore, we observed a surprising longevity of SARS-CoV-2 nucleic acid in saliva as a medium, across a range of storage temperatures, and after multiple freeze-thaws. We have demonstrated that the UoNATS assay is robust against what are inarguably highly heterogenous specimens and the potential inhibitory factors present within, with even the worst example we have encountered yielding a co-efficient of variation (CV%) of just 5.4%.

The technical validation of our in-house method was designed to address the required criteria for national UK testing standards, as defined by the guidance provided by UK regulatory and governing bodies; MHRA-TPP, Department for Health and Social Care (DHSC), and PHE. In 2021, the UoNATS service and testing protocol was assessed by UKAS for concordance with the requirements established by the International Standards Organisation for medical testing laboratories (ISO 17025:2017) and was duly awarded the status of a nationally accredited SARS-CoV-2 testing laboratory.

When reviewing the clinical concordance of the UoNATS assay with that of independent validated test providers, we observed positive and negative results concordance of 99.4% and 99.6%, respectively. We have presented data to show that the assay is both highly repeatable and reproducible. As per the quality standard limits defined by MHRA/PHE guidance ^[20]^, the required sensitivity for nationally approved SARS-CoV-2 diagnostic tests in the UK is the ability to detect down to 1 vc µL^-1^ of specimen, we confidently detected this viral load with 100% occurrence. The dynamic range of the assay was quantified and demonstrates that the assay maintains accuracy and precision from high (CT 15) to low (CT 40) viral loads.

Some saliva-based methods published in the recent literature incorporate the addition of extraneous regents such as Tween-20 or Tris-Borate-EDTA (TBE) to optimise molecular detection ^[25]^. We have found these additives to be unnecessary when used in combination with the methods presented here. For instance, the Quantabio ToughMix PCR mastermix is optimised for use with crude biological samples and resistant to potential PCR inhibitors associated with such samples. We have submitted our detailed methods and protocols in a separate publication ^[21]^ for any who wish to reproduce what we are doing.

Currently, the variants of concern are Omicron (B.1.1.529) and sub lineages, originating in South Africa. These are the dominant strains globally, representing between 95.4% and 99.3% of new sequences submitted in the last 30 days as of 18^th^ February 2022 (EpiCov; GISAID). Despite a prevalent C>T mismatch in the Charité-E primer region, we observe no evidence of detriment to PCR performance as a result of these variants. In the UK, national restrictions such as mandatory face coverings, social distancing, and lockdowns are ending however the prevalence of SARS-CoV-2 has not diminished; it is likely that sustained testing will be beneficial for some time still to come, particularly for the detection of new strains with the potential to avoid natural and induced immunity.

## Conclusion

The validation work we present here aimed to assess the performance of the UoNATS assay for the intended purpose as a SARS-CoV-2 nucleic acid detection test, as well as validating saliva as an appropriate specimen on which to determine infection status and make a COVID-19 diagnoses. The UoNATS assay can perform to the high specifications required for diagnostic SARS-CoV-2 RT-qPCR testing in the general population; meeting the performance criteria stipulated by UK governing bodies. Our assay is accurate and robust, with high sensitivity and specificity. Non-invasive saliva-based diagnostic tests have enormous potential to sustain surveillance of SARS-CoV-2.

## Data Availability

All data produced in the present study are available upon reasonable request to the authors

## Acknowledgements

With thanks to all the team members not directly involved in this manuscript; Paulina Durczak, Christopher McCusker, Damiano Spadoni, Bethan Roberts, Joseph Chappell, James Hassall, and to the wider UoNATS testing service staff. This research was funded by the University of Nottingham and Medical Research Council grant number MC_PC_20027.

## Author contributions

Experimental work was performed by: H.H.J., A.A.T.L., H.T., F.S.T., D.S., K.L-S., R.H., and T.B.; Manuscript preparation was performed by: H.H.J., A.A.T.L., I.L., F.S.T.; Supervision was provided by: C.D., C.S., and A.V.B.

## Data availability statement

Data available upon request from the corresponding authors.

