## Supplemental data for "Performance evaluation of a non-invasive one-step multiplex RT-qPCR assay for detection of SARS-CoV-2 direct from human saliva"

**Supplementary Information**

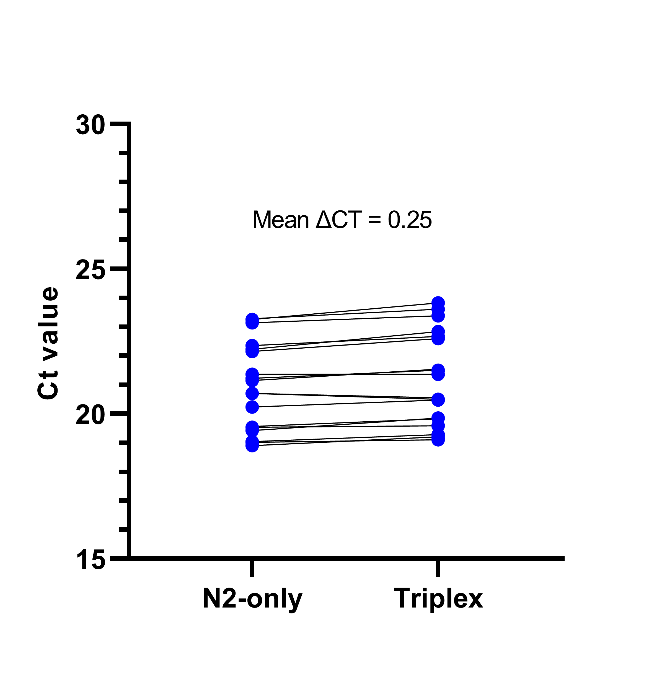

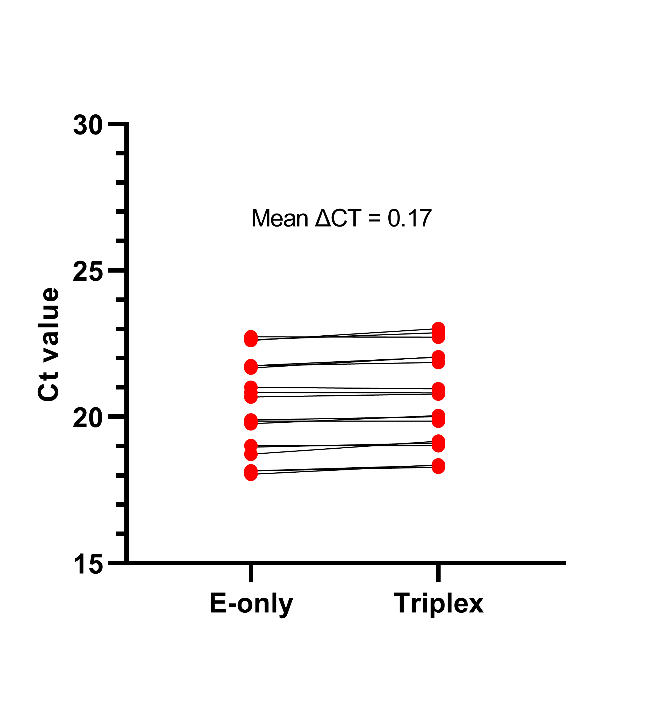

|  | *N2-only* | | | *Multiplex* | | | | **∆CT** | Mean ∆CT | *E-only* | | | | *Multiplex* | | | | **∆CT** | Mean ∆CT |
| --- | --- | --- | --- | --- | --- | --- | --- | --- | --- | --- | --- | --- | --- | --- | --- | --- | --- | --- | --- |
| **Standard** | **CT** | **Mean** | **SD** | **CT** | **Mean** | | **SD** |  |  | **CT** | **Mean** | | **SD** | **CT** | **Mean** | | **SD** |  |  |
| **1** | 19.02 | 18.98 | 0.078 | 19.11 | 19.2 | 0.087 | | 0.09 | 0.21 | 18.15 | 18.11 | 0.064 | | 18.28 | 18.33 | 0.046 | | 0.13 | 0.21 |
|  | 18.9 |  |  | 19.21 |  |  |  | 0.31 |  | 18.15 |  |  |  | 18.34 |  |  |  | 0.19 |  |
|  | 19.04 |  |  | 19.28 |  |  |  | 0.24 |  | 18.04 |  |  |  | 18.36 |  |  |  | 0.32 |  |
| **2** | 19.54 | 19.51 | 0.075 | 19.59 | 19.76 | 0.147 | | 0.05 | 0.25 | 19.02 | 18.91 | 0.155 | | 19.02 | 19.1 | 0.076 | | 0 | 0.19 |
|  | 19.42 |  |  | 19.86 |  |  |  | 0.44 |  | 18.73 |  |  |  | 19.17 |  |  |  | 0.44 |  |
|  | 19.56 |  |  | 19.83 |  |  |  | 0.27 |  | 18.97 |  |  |  | 19.11 |  |  |  | 0.14 |  |
| **3** | 20.7 | 20.55 | 0.273 | 20.5 | 20.51 | 0.04 | | -0.20 | -0.04 | 19.89 | 19.84 | 0.069 | | 20.01 | 19.96 | 0.103 | | 0.12 | 0.13 |
|  | 20.23 |  |  | 20.47 |  |  |  | 0.24 |  | 19.86 |  |  |  | 19.85 |  |  |  | -0.01 |  |
|  | 20.71 |  |  | 20.55 |  |  |  | -0.16 |  | 19.76 |  |  |  | 20.04 |  |  |  | 0.28 |  |
| **4** | 21.37 | 21.24 | 0.114 | 21.37 | 21.47 | 0.089 | | 0.00 | 0.23 | 20.68 | 20.84 | 0.165 | | 20.78 | 20.85 | 0.092 | | 0.1 | 0.01 |
|  | 21.22 |  |  | 21.51 |  |  |  | 0.29 |  | 20.83 |  |  |  | 20.82 |  |  |  | -0.01 |  |
|  | 21.15 |  |  | 21.54 |  |  |  | 0.39 |  | 21.01 |  |  |  | 20.96 |  |  |  | -0.05 |  |
| **5** | 22.15 | 22.24 | 0.099 | 22.6 | 22.7 | 0.117 | | 0.45 | 0.46 | 21.75 | 21.72 | 0.045 | | 21.86 | 21.99 | 0.107 | | 0.11 | 0.26 |
|  | 22.23 |  |  | 22.83 |  |  |  | 0.60 |  | 21.67 |  |  |  | 22.05 |  |  |  | 0.38 |  |
|  | 22.35 |  |  | 22.67 |  |  |  | 0.32 |  | 21.75 |  |  |  | 22.04 |  |  |  | 0.29 |  |
| **6** | 23.25 | 23.23 | 0.073 | 23.82 | 23.6 | 0.219 | | 0.57 | 0.38 | 22.61 | 22.66 | 0.071 | | 23.02 | 22.87 | 0.149 | | 0.41 | 0.21 |
|  | 23.14 |  |  | 23.38 |  |  |  | 0.24 |  | 22.74 |  |  |  | 22.72 |  |  |  | -0.02 |  |
|  | 23.28 |  |  | 23.61 |  |  |  | 0.33 |  | 22.63 |  |  |  | 22.87 |  |  |  | 0.24 |  |
|  |  |  |  |  |  | |  | 0.25 | |  |  | |  |  |  | |  | 0.17 | |

**Supplementary Figure & Table S1:** Comparison between single-target assays and the UoNATS triplex method. A clinical positive saliva sample was serially diluted and each of the resulting standards tested in PCR; either using individual assays in simplex or combined in the UoNATS triplex. Multiplexing the primer and probe assays in a single reaction resulted in minimal inhibition as seen by change of CT values between mono- and triplex of 0.25 for CDC-N2, and 0.17 for Charité-E.

**Supplementary Table S2:** Cross reactivity of the UoNATS assay to 21 respiratory organisms.

| Organism | Strain | CDC-N2 | Charité-E |
| --- | --- | --- | --- |
| *Adenovirus 1* | - | - | - |
| *Adenovirus 3* | N/A | - | - |
| *Adenovirus 31* | N/A | - | - |
| *B. parapertussis* | A747 | - | - |
| *B. pertussis* | A639 | - | - |
| *C. pneumoniae* | CWL-029 | - | - |
| *Coronavirus 229E* | N/A | - | - |
| *Coronavirus HKU-1* | Recombinant1 | - | - |
| *Coronavirus NL63* | N/A | - | - |
| *Coronavirus OC43* | N/A | - | - |
| *Influenza A H1N1pdm* | A/NY/02/092 | - | - |
| *Influenza AH1* | A/New Caledonia/20/99 | - | - |
| *Influenza AH3* | A/Brisbane/10/07 | - | - |
| *Influenza B* | B/Florida/02/06 | - | - |
| *M. pneumoniae* | M129 | - | - |
| *Metapneumovirus 8* | Peru6-20033 | - | - |
| *Parainfluenza 1* | N/A | - | - |
| *Parainfluenza 2* | N/A | - | - |
| *Parainfluenza 3* | N/A | - | - |
| *Parainfluenza 4* | N/A | - | - |
| *Rhinovirus 1A* | N/A | - | - |
| RSV A | N/A | - | - |
| SARS-CoV-2 | USA-WA1/20204 | **POSITIVE** | **POSITIVE** |
| Negative Control | A549 Alveolar cells | - | - |

**Supplementary Table S3 (a) and (b):** Prevalence of SARS-CoV-2 strains globally and in England, with mutations in the UoNATS target assay regions. The assays should withstand up to 2 mismatches and retain functionality. Mutation in the 3’ end of a sequence is prone to disrupt PCR performance and of higher concern.

**(a)**

| **Assay** | **Global strains with 2+ mutations in assay regions** | **Of which in the 3’ end** | **National strains with 2+ mutations in the assay regions** | **Of which in the 3’ end** | **National Strains with single mismatches** | **Of which in the 3’ end** |
| --- | --- | --- | --- | --- | --- | --- |
| US-CDC-N2 | 0.002% | 0.001% | 0 % | - | 1.065% | 0.217% |
| Charité-E | 0.008% | 0.000004% | 0 % | - | 0.227% | 0.049% |
|  | 765,421 sequences globally from 30/12/2019 to 8/06/2021 | | 149,964 sequences from England  from 30/12/2019 to 8/06/2021 | | | |

**(b)**

| **Omicron Lineage** | **Assay** | **National strains with 2+ mutations in each assay region** | **National strains with 1+ mutations in the 3’ ends** |
| --- | --- | --- | --- |
| B.1.1.529 | US-CDC-N2 | 0/95 | 0/95 |
|  | Charité-E | 0/95 | 0/95 |
| BA.1 | US-CDC-N2 | 0/100 | 0/100 |
|  | Charité-E | 0/100 | 0/100 |
| BA1.1 | US-CDC-N2 | 0/100 | 0/100 |
|  | Charité-E | 0/100 | 0/100 |
| BA.2 | US-CDC-N2 | 0/100 | 1/100 |
|  | Charité-E | 0/100 | 0/100 |
| BA.3 | US-CDC-N2 | 0/100 | 0/100 |
|  | Charité-E | 0/100 | 0/100 |
| **Total Sequences** | US-CDC-N2 | 0/495 | 1/495 (0.2%) |
|  | Charité-E | 0/495 | 0/495 |

**Supplementary Table S4:** Sample stability over 20 days under various storage conditions

| Sample 1 | | CDC-N2 | | | | Charité-E | | |
| --- | --- | --- | --- | --- | --- | --- | --- | --- |
|  | RT | | 4 ᵒC | -80 ᵒC | RT | | 4 ᵒC | -80 ᵒC |
| Number of tests | 10 | | 10 | 8 | 10 | | 10 | 8 |
| Mean CT | 30.19 | | 30.19 | 29.75 | 29.63 | | 29.64 | 29.29 |
| Std. Deviation | 0.2663 | | 0.3261 | 0.2497 | 0.3929 | | 0.4395 | 0.3605 |
| Coefficient of variation | 0.882% | | 1.080% | 0.839% | 1.326% | | 1.483% | 1.231% |

| Sample 2 | | CDC-N2 | | | | Charité-E | | |
| --- | --- | --- | --- | --- | --- | --- | --- | --- |
|  | RT | | 4 ᵒC | -80 ᵒC | RT | | 4 ᵒC | -80 ᵒC |
| Number of tests | 10 | | 10 | 8 | 10 | | 10 | 8 |
| Mean CT | 24.89 | | 24.92 | 24.93 | 23.94 | | 23.98 | 24.10 |
| Std. Deviation | 0.3973 | | 0.4038 | 0.3039 | 0.4550 | | 0.5202 | 0.3306 |
| Coefficient of variation | 1.597% | | 1.620% | 1.219% | 1.901% | | 2.169% | 1.371% |

| Sample 3 | CDC-N2 | | | | Charité-E | |
| --- | --- | --- | --- | --- | --- | --- |
|  | RT | 4 ᵒC | -80 ᵒC | RT | 4 ᵒC | -80 ᵒC |
| Number of tests | 10 | 10 | 8 | 10 | 10 | 8 |
| Mean CT | 27.77 | 27.12 | 26.93 | 27.28 | 26.69 | 26.56 |
| Std. Deviation | 0.6691 | 0.2931 | 0.3507 | 0.4848 | 0.4226 | 0.4408 |
| Coefficient of variation | 2.410% | 1.081% | 1.302% | 1.777% | 1.583% | 1.659% |

**Supplementary Table S5:** Descriptive statistics for the repeated measurements of a saliva sample at the limit of detection tested in multiple technical replicates.

|  | **CDC-N2** | **Charité-E** |
| --- | --- | --- |
| Number of Replicates | 23 | 24 |
| Mean CT | 36.00 | 34.38 |
| Standard Deviation | 0.9333 | 1.008 |
| Coefficient of variation | 2.593% | 2.932% |

**Supplementary Table S****6:** Descriptive statistics for the reproducibility of the 2 UoNATS assay targets N2 and E. Seven SARS-CoV-2 positive samples were tested separately in triplicate by four individual operators using different reagents and equipment. The variation in CT is shown to be low and within acceptable ranges. Sample 7 (*) was included to exemplify a problematic sample. The sample had a thick gel-like consistency and was difficult to dispense 8 µL into the reaction; however, the coefficient of variation was still below 5.4%.

| **CDC-N2 (FAM)** | | **Sample 1** | **Sample 2** | **Sample 3** | **Sample 4** | **Sample 5** | **Sample 6** | **Sample 7 *** |
| --- | --- | --- | --- | --- | --- | --- | --- | --- |
| Replicates | | 12 | 12 | 12 | 11 | 12 | 12 | 12 |
| Mean CT | | 27.95 | 23.73 | 23.26 | 18.06 | 26.51 | 29.64 | 19.10 |
| Std. Deviation | | 0.220 | 0.226 | 0.243 | 0.253 | 0.500 | 0.556 | 1.024 |
| Coefficient of variation | | 0.788% | 0.952% | 1.044% | 1.403% | 1.886% | 1.876% | 5.362% |
| **Charité-E (HEX)** | | **Sample 1** | **Sample 2** | **Sample 3** | **Sample 4** | **Sample 5** | **Sample 6** | **Sample 7 *** |
| Replicates | | 12 | 12 | 12 | 11 | 12 | 12 | 12 |
| Mean CT | | 27.82 | 23.53 | 23.39 | 18.21 | 26.10 | 29.40 | 18.90 |
| Std. Deviation | | 0.155 | 0.202 | 0.219 | 0.095 | 0.425 | 0.483 | 0.988 |
| Coefficient of variation | | 0.559% | 0.858% | 0.938% | 0.521% | 1.627% | 1.643% | 5.227% |

**Supplementary Table S7:** Calculation of positive and negative predictive value for first-line screening and second confirmatory test

| **Initial Screening** |  |  | **True Positives** | **True negatives** | **False Negatives** | **False positives** |  |  |
| --- | --- | --- | --- | --- | --- | --- | --- | --- |
| **Incidence considered** (Population size of 10,000) | Total positive cases  =  Population * incidence | Total negative cases  =  Population * (100% - incidence) | Positives detected = Positive cases * Diagnostic Sensitivity | Negatives detected = Negative cases * Diagnostic Specificity | Positives missed =  Positive cases * (100%-Diag. Sensitivity) | Negatives missed =  Negative cases * (100%-Diag. Specificity) | **Positive Predictive Value  = true positives detected / ALL positive results** | **Negative Predictive Value  = true negatives detected / ALL negative results** |
| 0.61% | 61 | 9939 | 61 | 9899 | 0.4 | 40 | 60% | 100% |
| 1% | 100 | 9900 | 99 | 9860 | 0.6 | 40 | 72% | 100% |
| 5% | 500 | 9500 | 497 | 9462 | 3 | 38 | 93% | 100% |
| 10% | 1000 | 9000 | 994 | 8964 | 6 | 36 | 97% | 100% |
| 25% | 2500 | 7500 | 2485 | 7470 | 15 | 30 | 99% | 100% |
| 50% | 5000 | 5000 | 4970 | 4980 | 30 | 20 | 100% | 99% |
| **Confirmatory Test** |  |  | **True Positives** | **True negatives** | **False Negatives** | **False positives** |  |  |
| **Incidence considered** (Population = number of repeat tests) | Total positive cases  =  Population * incidence | Total negative cases  =  Population * (100% - incidence) | Positives detected = Positive cases * Diagnostic Sensitivity | Negatives detected = Negative cases * Diagnostic Specificity | Positives missed =  Positive cases * (100%-Diag. Sensitivity) | Negatives missed =  Negative cases * (100%-Diag. Specificity) | **Positive Predictive Value  = true positives detected / ALL positive results** | **Negative Predictive Value  = true negatives detected / ALL negative results** |
| 0.61% | 0.6 | 99.8 | 0.6 | 99 | 0.00 | 0.4 | 99.4% | 100% |
| 1% | 1.4 | 137.6 | 1.4 | 137 | 0.01 | 0.6 | 99.5% | 100% |
| 5% | 26.8 | 508.3 | 26.6 | 506 | 0.16 | 2.0 | 100% | 100% |
| 10% | 103.0 | 927.0 | 102.4 | 923 | 0.62 | 3.7 | 100% | 100% |
| 25% | 628.8 | 1886.3 | 625.0 | 1879 | 3.77 | 7.5 | 100% | 100% |
| 50% | 2495.0 | 2495.0 | 2480.0 | 2485 | 14.97 | 10.0 | 100% | 100% |
